# Unintended pregnancy among women in Nepal: Socioeconomic, fertility, and contraceptive determinants from the 2022 NDHS

**DOI:** 10.1101/2025.08.27.25334549

**Authors:** Ashraful Alam Siddique, Towhida Nasrin, Maliha Mehzabeen, Orin Akter, Md. Golam Rabbani, Shehrin Shaila Mahmood

**Affiliations:** Health Systems and Population Studies Division, icddr,b, Dhaka, Bangladesh; Maternal and Child health Division, icddr,b, Dhaka, Bangladesh; Power and Participation Research Centre – PPRC, Dhaka, Bangladesh

**Keywords:** Unintended pregnancy, unmet contraceptive need, reproductive health, family planning, DHS, Nepal

## Abstract

**Background:** Globally, nearly half of all pregnancies are unintended, with a disproportionately high occurrence in LMIC countries. Unintended pregnancies present significant public health challenges, leading to an elevated risk of unsafe abortions, maternal mortality, and substantial economic burdens. While Nepal has made progress in reducing its unintended pregnancy rate over the past three decades, the high rate of abortion for unintended pregnancies remains a serious concern. This study examines the association of unintended pregnancies among Nepalese women with socioeconomic, fertility, and contraceptive factors, providing evidence to guide policy and interventions for improved reproductive health outcomes.

**Methods:** Utilizing weighted data from the 2022 Nepal Demographic and Health Survey (NDHS), this cross-sectional study included 2,992 married women aged 15– 49. The primary outcome was unintended pregnancy, while the main exploratory variable was unmet need for contraception, determined by a DHS-endorsed algorithm. We also examined various socio-demographic and reproductive factors, including age, education, wealth, and desire for more children. The data were analyzed using descriptive statistics, bivariate analysis (chi-square test), and multivariable logistic regression to identify significant predictors.

**Results:** The study found a prevalence of 27.9% for unintended pregnancy and 29.4% for unmet need for contraception. The highest rates of unintended pregnancies were observed in women aged 35–49 and 15–19, as well as in those with lower education and wealth levels. Multivariable logistic regression revealed that having an unmet need for contraception was the strongest predictor (aOR = 14.05, 95% CI = 6.14– 14.89). Other significant factors included current use of contraception (aOR = 9.56, 95% CI = 6.14–14.89), not desiring more children (aOR = 2.28, 95% CI = 1.83– 2.83), being in the poorest wealth quintile, and having a husband with only primary education. Regional disparities were also evident, with Karnali Province reporting the highest rate of unintended pregnancies (38%).

**Conclusions:** This study establishes a strong link between unmet contraceptive needs and unintended pregnancies in Nepal. The findings highlight the critical importance of improving access to family planning services and addressing systemic barriers to consistent contraceptive use. Interventions should be tailored to vulnerable subgroups, particularly adolescents, older women, and those in the poorest wealth quintiles. A comprehensive approach that supports informed reproductive choices is essential for reducing unintended pregnancies and advancing reproductive health outcomes in Nepal.

**WHAT IS ALREADY KNOWN ON THIS TOPIC:**

⇒ Nearly half of all pregnancies worldwide are unintended, with a higher burden in low- and middle-income countries.
⇒ Unmet need for contraception is a major contributor to unintended pregnancies.
⇒ Evidence on the direct link between unmet contraceptive need and unintended pregnancy in South Asia, especially Nepal, is limited.
⇒ Nepal has reduced unintended pregnancy rates overall, but abortions among unintended pregnancies have increased, and unmet need for contraception remains high.

**WHAT THIS STUDY ADDS:**

⇒ Shows a strong, independent association between unmet need for contraception and unintended pregnancy in Nepal.
⇒ Women with unmet need were over 14 times more likely to report unintended pregnancy than those without unmet need.
⇒ Identifies key socio-demographic predictors: younger age, lower wealth, limited husband’s education, and completed fertility preferences.
⇒ Reveals significant provincial disparities in unintended pregnancy rates across Nepal.
⇒ Highlights systemic barriers to effective and consistent contraceptive access despite national family planning efforts.

**HOW THIS STUDY MIGHT AFFECT RESEARCH, PRACTICE OR POLICY:**

⇒ Provides robust evidence to guide reproductive health and family planning policy in Nepal.
⇒ Supports strengthening FP-2030 commitments with a focus on underserved populations.
⇒ Calls for ensuring consistent availability of modern contraceptives at all levels of the health system.
⇒ Recommends integrating culturally sensitive counseling, adolescent-focused reproductive health education, and male partner involvement.
⇒ Suggests future research should explore socio-cultural barriers through mixed-methods approaches to inform targeted interventions.

## BACKGROUND

Approximately half of all pregnancies are unintended across the world (1), with a higher proportion occurring in low- and middle-income countries (2). Unintended pregnancies present a major public health challenge, often leading to unsafe abortion, maternal mortality, and long-term health and economic burden (1). According to recent evidence, almost 30% of all pregnancies are terminated, a number that rises beyond an alarming 60% threshold when looking at unintended pregnancies. Alarmingly, half of these abortions are unsafe, leading to serious health complications and contributing substantially to maternal mortality (33). Each year, around 7 million women require hospitalization due to complications from these procedures, and this contributes to 5-13% of all maternal deaths, making abortion a primary reason for maternal mortality. On the other hand, the economic burden of treating complications from unsafe abortions in developing countries is estimated to be $553 million per year (1). Between 2015 and 2019, unintended pregnancy rates showed significant regional variations. In Europe and North America, the annual unintended pregnancy rate was reported to be 35 per 1000 women aged between 15 and 49 years. In contrast, Central and Southern Asia experienced a higher rate of 64 unintended pregnancies per 1,000 women. Sub-Saharan Africa faced the highest incidence, with 91 unintended pregnancies annually per 1,000 women in the same age group (1). Meanwhile, Nepal, one of the poorest and slowest-growing economies in South Asia, has seen significant changes in its unintended pregnancy rates (3). Between 2015 and 2019, unintended pregnancy rates showed wide regional variation: 35 per 1,000 women in Europe and North America, 64 in Central and Southern Asia, and 91 in Sub-Saharan Africa. In Nepal, unintended pregnancy rates have declined by 45% between 1990–1994 and 2015–2019, but the abortion rate for unintended pregnancies increased dramatically from 34% to 69% during the same period, which is a major concern (4). One of the key goals of the agenda is ensuring gender equality, as it is considered a pillar of human development. An indicator of gender equality is stated to be “the proportion of women aged 15 to 49 years who make their own informed decisions regarding sexual relations, contraceptive use, and reproductive health care” (5). The most recent data on this (SDG 5.6.1), which looked at partnered women of reproductive age in 64 countries, found that 23% are unable to refuse sex, 24% are unable to make choices regarding their own health care, and 8% are unable to make decisions specifically about contraception, indicating that a significant proportion of women are unable to exercise decision-making power over their sexual and reproductive health and rights (1). To address this issue, it is essential to identify the factors correlated with unintended pregnancy (35). Several such factors contributing to unintended pregnancies have been identified, including limited access to reproductive health services, high rates of contraceptive discontinuation, uncertainty about fertility goals, and instances of gender-based violence (6,7). Yet the link between unmet contraceptive needs and unintended pregnancies remains inadequately explored. Despite efforts over the past two decades to improve reproductive health outcomes, Nepal has a maternal mortality rate of 151 per 100,000 live births (8). While the unmet need for contraception has shown a slight decrease from 24.7% in 2006 to 23.7% in 2016 (9), the unintended pregnancy rate remains high at 68 per 1,000 women of reproductive age (10). Evidence on the association between unmet needs and unintended pregnancies in the context of Nepal remains limited. Understanding the factors underlying these pregnancies, particularly the role of unmet needs for contraception, is crucial for developing effective interventions.

This study aimed to bridge the gap in current research exploring how unmet contraception, along with socioeconomic and fertility-related factors, influences unintended pregnancies among Nepalese women. Insights from this analysis can help policymakers and healthcare providers develop targeted strategies to improve reproductive health outcomes and support informed family planning decisions in Nepal.

## METHODS

### Data source, study design, and study population

This study was conducted utilizing the dataset of Nepal Demographic and Health Survey (NDHS) 2022, carried out by the Department of Health Services, Ministry of Health and Population in Nepal (11). A cross-sectional design was implemented to survey married women between the ages of 15 and 49, providing a nationally representative sample. A total of 14,845 women from that age group were successfully interviewed in 14,845 households from 476 enumeration areas (EA) (248 in urban and 228 in rural areas) in all seven Nepalese provinces. Provinces were stratified into urban and rural areas, with EAs selected using probability proportional to size (PPS). Response rates were 97% overall and 95% for the specific group. The study population was 2,992 women who were extracted from 14,845 based on the study criterion. Details of the survey were publicly available on “The DHS Program” website, with technical support provided by UNICEF, ICF International through the Demographic and Health Surveys (DHS) Program, and the World Health Organization (WHO).

### Measurement of variables

#### Outcome variable

In this study, we recorded the pregnancy intention status of women regarding their most recent pregnancy as the outcome variable. For the analysis, we considered only the most recent completed pregnancy within the preceding five years. Current pregnancies were excluded from the study. The participants were asked if their last successful pregnancy was wanted, and they could answer with wanted, wanted later, and not wanted. For this analysis, we considered wanted later and not wanted as the negative response while wanting the pregnancy obviously indicated an affirmative response.

### Exploratory variables

The major exploratory variable for this study was the unmet need for contraception. Contraception utilization status was recorded for participants regardless of their reported desire for spacing or limiting childbirth. Unmet need for contraception was not based on direct responses from participants. Instead, it was determined using the DHS-endorsed algorithm developed by Bradley (29). This method uses a combination of indicators such as fertility preferences, contraceptive use status, fecundity, current pregnancy status, and sexual activity to assess need. Women were considered to have an unmet need if they were sexually active, biologically able to conceive, not using any form of contraception, and either wished to delay childbearing for at least two years (spacing) or wanted no more children (limiting). For analysis purposes, we recoded the outcome into two categories: “unmet need” (which includes both spacing and limiting) and “no unmet need” (including those using contraception or not classified as having unmet need). This approach is consistent with global standards for measuring reproductive health indicators. For the analysis, we combined these categories into two groups: “Yes” for any unmet need (unmet need for limiting or spacing) and “No” for no unmet need or use of contraception for either limiting or spacing. The analysis excluded women who were unmarried, without a sexual history, infecund, sterilized (themselves or their partner), or planning to have a child within two years. Based on the relevant literature review, we selected some covariates, and according to the following criteria for this analysis. We categorized the age of the study population into five groups as follows: 15-19, 20-24, 25-29, 30-34, and 35-49. Residency was classified as either Urban or Rural. Education levels were categorized as ‘No Education’, ‘Primary’, ‘Secondary’, or ‘Higher’. The husband’s education was similarly classified. Participants’ religion was recorded as Hindu, Buddhist, Muslim, or Other. The household head was identified as either Male or Female. Employment status was noted as currently working (Yes) or not working (No). The respondent’s occupation was categorized into ‘Not working’, ‘Agriculture’, ‘Services’, ‘Sales’, ‘Skilled’, or ‘Unskilled’. The desire for more children was recorded as Yes or No. Decision-making authority regarding the respondent’s healthcare was also noted, categorized as ‘Yes’ or ‘No’. Whether the respondent had ever experienced a terminated pregnancy was recorded as ‘Yes’ or ‘No’. Lastly, the use of any contraception at the time of the survey was documented as ‘Yes’ or ‘No’. The wealth index or socioeconomic status in the DHS was determined using principal component analysis (PCA) into five groups: poorest, poor, middle, rich, and richest based on household assets and ownership (**Table 1**).

**Table 1:**
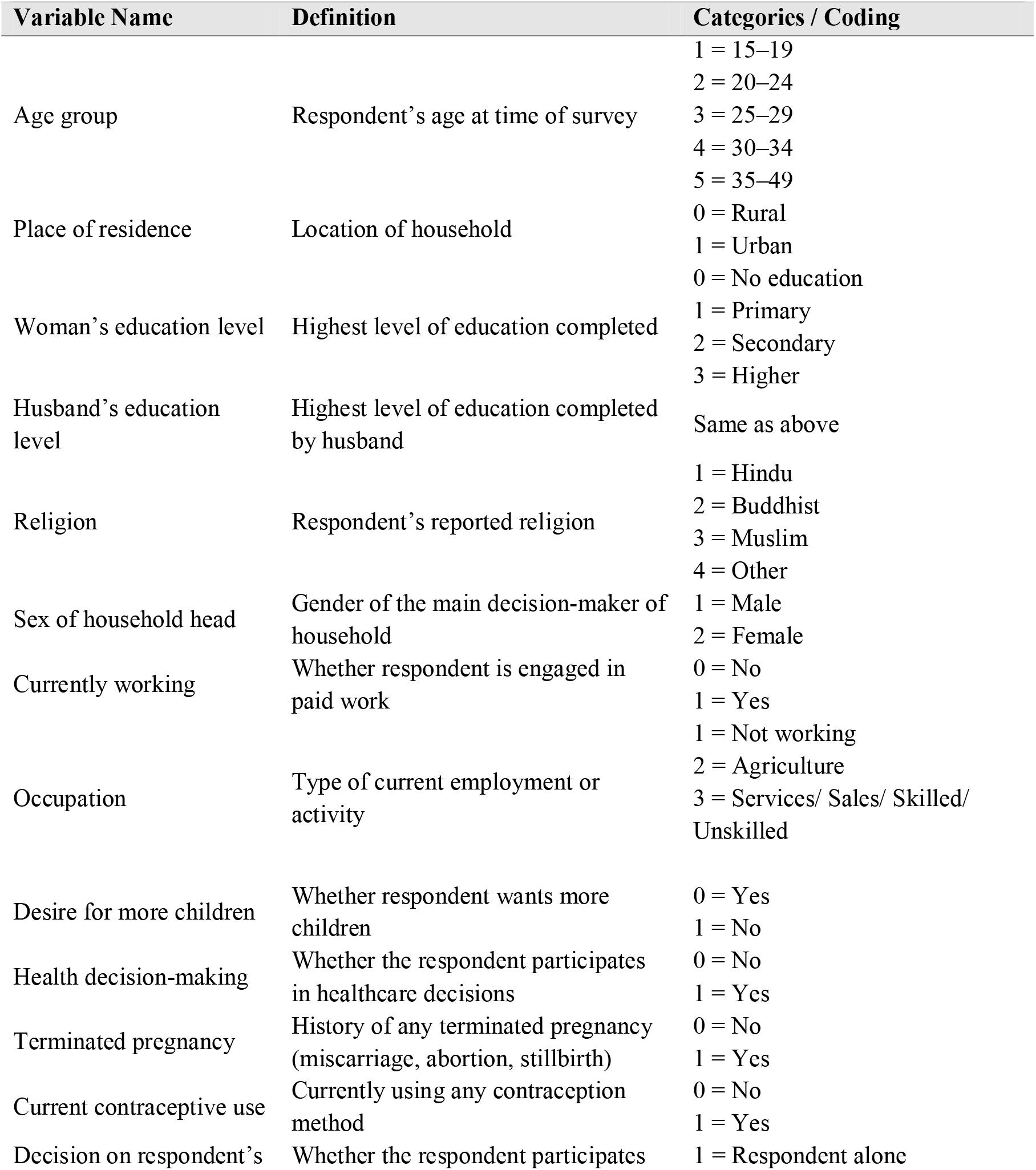
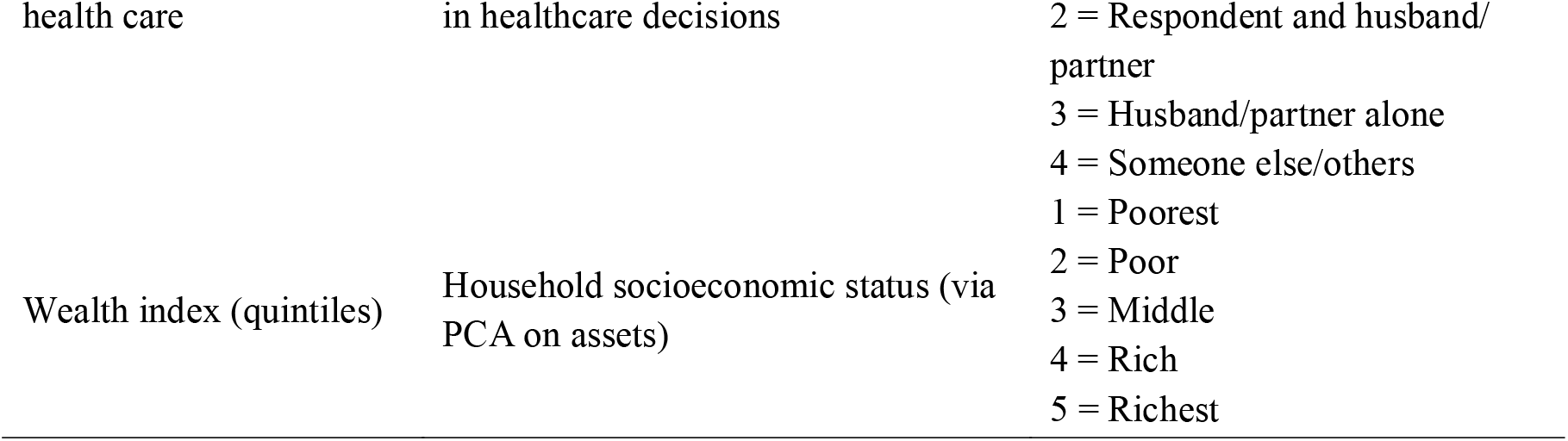
Description and coding of covariates used in the analysis.

### Data analysis

In this study, data were analyzed using Stata-16. We used the weighted sample in this analysis. Additionally, we performed descriptive, bivariate, and multivariate analyses. The utilization of descriptive analysis allowed for the exploration of socio-demographic characteristics and their distributions. Meanwhile, bivariate analysis (chi-square test) was utilized to identify the association between unintended pregnancy and exploratory variables. Then, multicollinearity was checked for the exploratory variables so that further analysis could be performed. Multivariable logistic regression analysis, which was performed to quantify the effect of an explanatory variable on the outcome variable, was only done with variables that showed statistically significant association (p<0.05) in the chi-square test. Results were presented as odds ratios with 95% confidence intervals. All tests were considered significant at the 5% level. The tests were conducted at the 5% significance level, and the odds ratios were expressed with 95% confidence intervals.

### Results

The prevalence of unintended pregnancy among women in the study was 27.9%, while 29.4% of women reported an unmet need for contraception (**Figure 1**).

**Figure 1.**
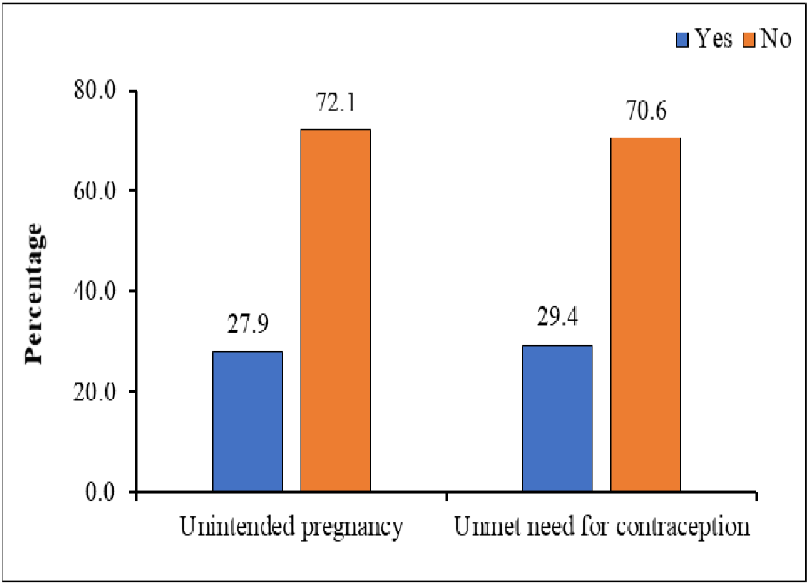
Prevalence of unintended pregnancy and unmet need for contraception among women in Nepal, 2022

**Table 2** presents the socio-demographic and reproductive characteristics of the 2,992 women of reproductive age included in this analysis, based on weighted data from the 2022 Nepal Demographic and Health Survey (NDHS). The majority of participants were aged between 20–29 years, with about one-third (33.9%) aged 20–24 and another 31.8% aged 25–29. Two-thirds (65.9%) of the women lived in urban areas. Most had received at least secondary education, and over 70% of their husbands had completed secondary or higher education. Nearly 70% of households were headed by men. About half of the women were currently working, with the largest group engaged in agriculture.

**Table 2.**
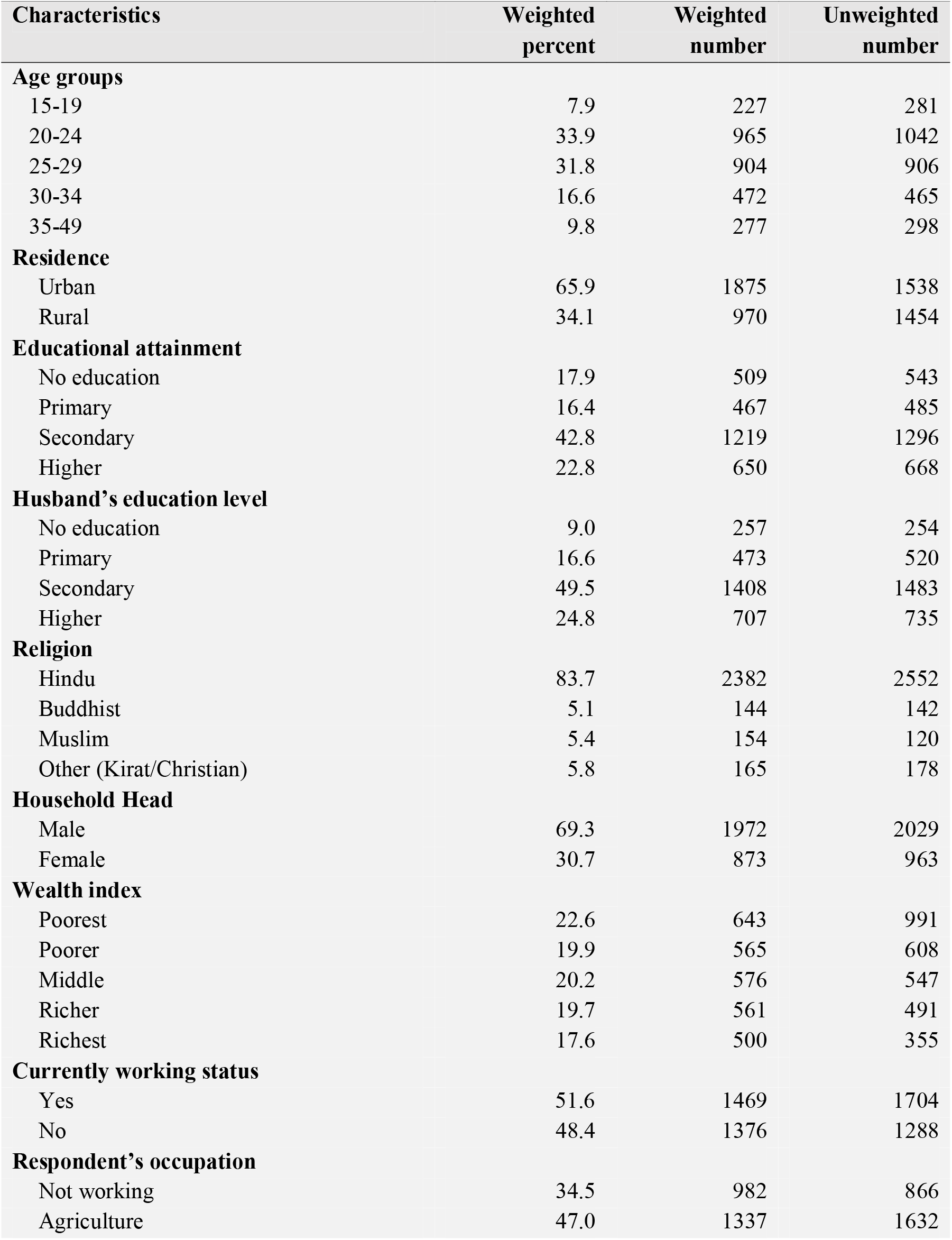
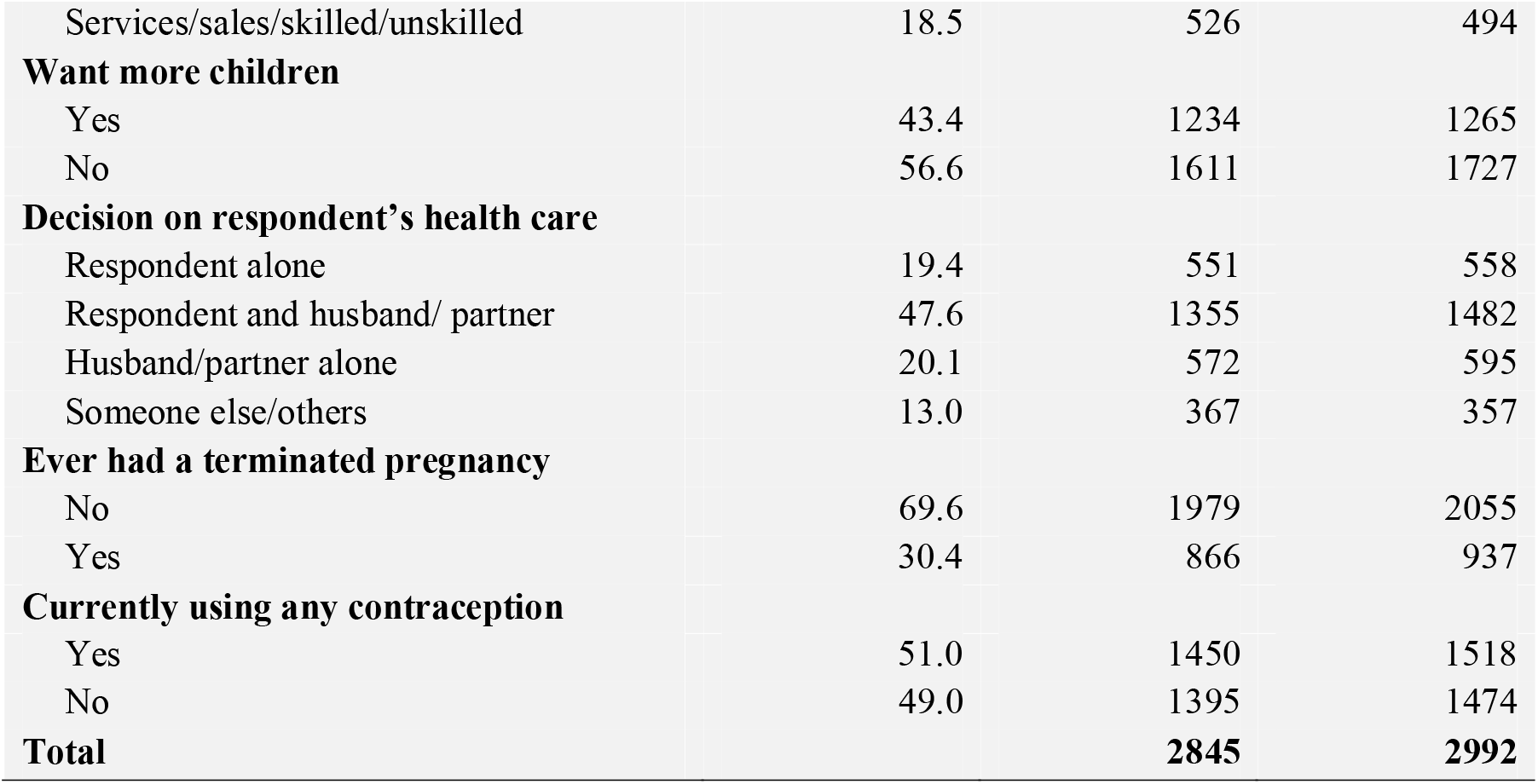
Background characteristics of the sample, NDHS-2022.

| Characteristics | Weighted percent | Weighted number | Unweighted number |
| --- | --- | --- | --- |
| **Age groups** |  |  |  |
| 15-19 | 7.9 | 227 | 281 |
| 20-24 | 33.9 | 965 | 1042 |
| 25-29 | 31.8 | 904 | 906 |
| 30-34 | 16.6 | 472 | 465 |
| 35-49 | 9.8 | 277 | 298 |
| **Residence** |  |  |  |
| Urban | 65.9 | 1875 | 1538 |
| Rural | 34.1 | 970 | 1454 |
| **Educational attainment** |  |  |  |
| No education | 17.9 | 509 | 543 |
| Primary | 16.4 | 467 | 485 |
| Secondary | 42.8 | 1219 | 1296 |
| Higher | 22.8 | 650 | 668 |
| **Husband's education level** |  |  |  |
| No education | 9.0 | 257 | 254 |
| Primary | 16.6 | 473 | 520 |
| Secondary | 49.5 | 1408 | 1483 |
| Higher | 24.8 | 707 | 735 |
| **Religion** |  |  |  |
| Hindu | 83.7 | 2382 | 2552 |
| Buddhist | 5.1 | 144 | 142 |
| Muslim | 5.4 | 154 | 120 |
| Other (Kirat/Christian) | 5.8 | 165 | 178 |
| **Household Head** |  |  |  |
| Male | 69.3 | 1972 | 2029 |
| Female | 30.7 | 873 | 963 |
| **Wealth index** |  |  |  |
| Poorest | 22.6 | 643 | 991 |
| Poorer | 19.9 | 565 | 608 |
| Middle | 20.2 | 576 | 547 |
| Richer | 19.7 | 561 | 491 |
| Richest | 17.6 | 500 | 355 |
| **Currently working status** |  |  |  |
| Yes | 51.6 | 1469 | 1704 |
| No | 48.4 | 1376 | 1288 |
| **Respondent's occupation** |  |  |  |
| Not working | 34.5 | 982 | 866 |
| Agriculture | 47.0 | 1337 | 1632 |
| Services/sales/skilled/unskilled | 18.5 | 526 | 494 |
| **Want more children** |  |  |  |
| Yes | 43.4 | 1234 | 1265 |
| No | 56.6 | 1611 | 1727 |
| **Decision on respondent's health care** |  |  |  |
| Respondent alone | 19.4 | 551 | 558 |
| Respondent and husband/ partner | 47.6 | 1355 | 1482 |
| Husband/partner alone | 20.1 | 572 | 595 |
| Someone else/others | 13.0 | 367 | 357 |
| **Ever had a terminated pregnancy** |  |  |  |
| No | 69.6 | 1979 | 2055 |
| Yes | 30.4 | 866 | 937 |
| **Currently using any contraception** |  |  |  |
| Yes | 51.0 | 1450 | 1518 |
| No | 49.0 | 1395 | 1474 |
| **Total** |  | **2845** | **2992** |

More than half of the participants (56.6%) said they did not want any more children, while just under half (51%) were using some form of contraception. Around 30% had experienced a terminated pregnancy.

Notably the regional differences reveal in pregnancy intention across Nepal’s provinces. Karnali Province reported the highest proportion of unintended pregnancies (38%), followed by Gandaki (31%) and Lumbini (30%). In contrast, Madhesh and Koshi Provinces had the lowest rates, at 25% and 26% respectively (**Figure 2)**. Karnali Province had the highest proportion of women who reported wanting no more children (20%), followed by Gandaki (18%) and Lumbini (13%).

**Figure 2.**
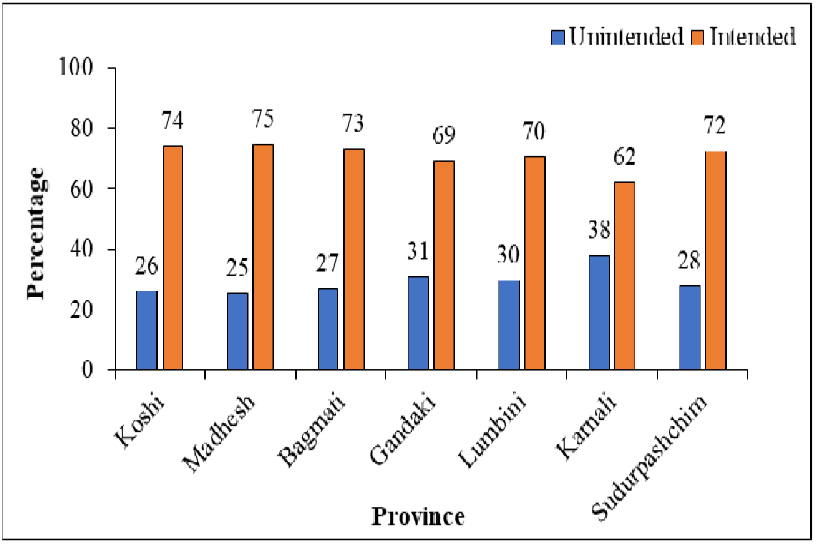
Regional variation in pregnancy intention among women in Nepal, 2022

In contrast, Madhesh had the lowest proportion of women expressing a desire to stop childbearing (8%), despite having a relatively high percentage (17%) of women who wanted to delay their next pregnancy (**Figure 3**).

**Figure 3.**
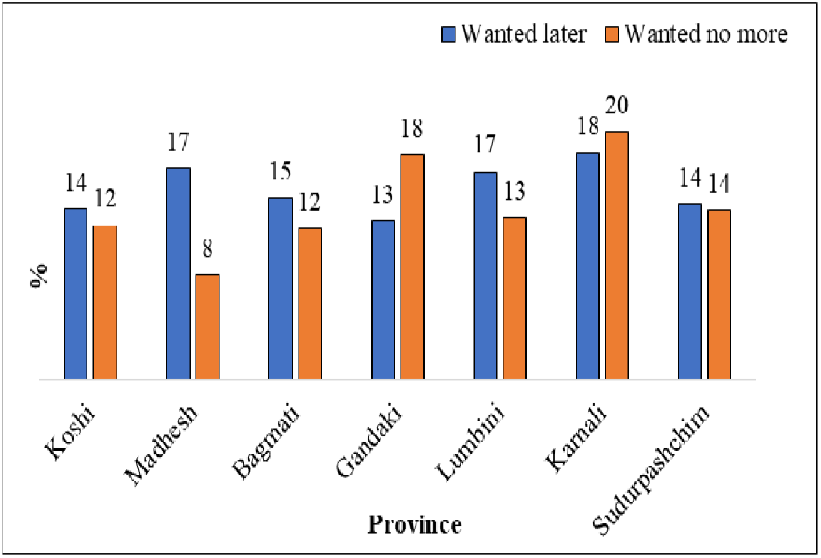
Type of unintended pregnancy by province among married women in Nepal, 2022

**Table 3** outlines the relationship between unintended pregnancy and various demographic and socio-economic variables. The data indicate significant age-related differences, with the highest levels of unintended pregnancies among women aged 35–49 (47.4%) and adolescents aged 15–19 (31.6%) (p < 0.001), suggesting that both younger and older women are particularly at risk. Education appears to play a protective role. Women with higher education had the lowest proportion of unintended pregnancies (24.2%), whereas those with no formal education experienced higher rates (31.1%) (p = 0.009). This inverse relationship also holds for the education level of husbands, where unintended pregnancy rates were lower among women whose husbands had higher education (p = 0.002). Economic status, as measured by the wealth index, was also significantly associated with unintended pregnancy. Women in the poorest quintile reported a higher prevalence (34.5%) compared to those in the richest group (22.7%) (p = 0.001). Additionally, women engaged in paid work had a greater incidence (30.7%) than those not currently working (24.9%) (p = 0.042), possibly pointing to work-related stressors or limited access to reproductive health services. Strong associations were also found between reproductive history and unintended pregnancy. Women who expressed a desire to have no more children were more likely to experience an unintended pregnancy (36.3%) than those who still wanted children (19.7%) (p < 0.001). Likewise, women who had ever had a pregnancy termination (45.5%) or reported unmet need for contraception (38.1%) were more likely to report their current or recent pregnancy as unintended (p < 0.001 for both), suggesting unmet needs in reproductive planning and contraceptive coverage.

**Table 3.** Demographic characteristics by unintended pregnancy, NDHS-2022.

| Characteristics | Unintended<br>(Weighted %) | Intended<br>(Weighted %) | p-value<sup>1</sup> |
| --- | --- | --- | --- |
| **Age groups** |  |  |  |
| 15-19 | 31.6 | 68.4 | 0.000 |
| 20-24 | 25.7 | 74.3 |  |
| 25-29 | 24.3 | 75.7 |  |
| 30-34 | 26.0 | 74.0 |  |
| 35-49 | 47.4 | 52.6 |  |
| **Residence** |  |  |  |
| Urban | 27.7 | 72.3 | 0.854 |
| Rural | 28.2 | 71.8 |  |
| **Educational attainment** |  |  |  |
| No education | 31.1 | 68.9 | 0.009 |
| Primary | 32.1 | 67.9 |  |
| Secondary | 26.9 | 73.1 |  |
| Higher | 24.2 | 75.8 |  |
| **Husband's education level** |  |  |  |
| No education | 26.1 | 73.9 | 0.002 |
| Primary | 35.9 | 64.1 |  |
| Secondary | 27.5 | 72.5 |  |
| Higher | 24.0 | 76.0 |  |
| **Religion** |  |  |  |
| Hindu | 28.6 | 71.4 | 0.029 |
| Buddhist | 21.2 | 78.8 |  |
| Muslim | 18.0 | 82.0 |  |
| Other (Kirat/Christian) | 32.9 | 67.1 |  |
| **Household Head** |  |  |  |
| Male | 27.0 | 73.0 | 0.342 |
| Female | 30.0 | 70.0 |  |
| **Wealth index** |  |  |  |
| Poorest | 34.5 | 65.5 | 0.001 |
| Poorer | 26.7 | 73.3 |  |
| Middle | 27.9 | 72.1 |  |
| Richer | 26.2 | 73.8 |  |
| Richest | 22.7 | 77.3 |  |
| **Currently working status** |  |  |  |
| Yes | 30.7 | 69.3 | 0.042 |
| No | 24.9 | 75.1 |  |
| **Respondent's occupation** |  |  |  |
| Not working | 26.9 | 73.1 | 0.069 |
| Agriculture | 29.5 | 70.5 |  |
| Services/sales/skilled/unskilled | 32.8 | 67.2 |  |
| **Want more children** |  |  |  |
| Yes | 19.7 | 80.3 | 0.000 |
| No | 36.3 | 63.7 |  |
| **Decision on respondent's health care** |  |  |  |
| Respondent alone | 32.3 | 67.7 | 0.095 |
| Respondent and husband/ partner | 26.4 | 73.6 |  |
| Husband/partner alone | 29.0 | 71.0 |  |
| Someone else/others | 25.1 | 74.9 |  |
| **Ever had a terminated pregnancy** |  |  |  |
| Yes | 45.5 | 54.5 | 0.000 |
| No | 20.2 | 79.8 |  |
| **Currently using any contraception** |  |  |  |
| Yes | 31.1 | 68.9 | 0.000 |
| No | 24.6 | 75.4 |  |
| **Unmet need** |  |  |  |
| Yes | 38.1 | 61.9 | 0.000 |
| No | 23.6 | 76.4 |  |
| <sup>1</sup>p-values from Pearson's chi-square tests for categorical comparisons; <sup>1</sup>p < 0.05 |  |  |  |

The data indicate a clear inverse relationship between wealth status and the prevalence of unintended pregnancy. Women in the poorest wealth quintile experienced the highest rate of unintended pregnancy at 35%, while those in the richest quintile had the lowest rate at 23%. The prevalence gradually decreased across the wealth categories, with poorer, middle, and richer groups reporting rates of 27%, 28%, and 26%, respectively **(Figure 4)**. Results of the logistic regression analysis showed the factors associated with unintended pregnancy. Having an unmet need for contraception emerged as the strongest predictor of reporting unintended pregnancy, with an odds ratio of 14.05.

**Figure 4.**
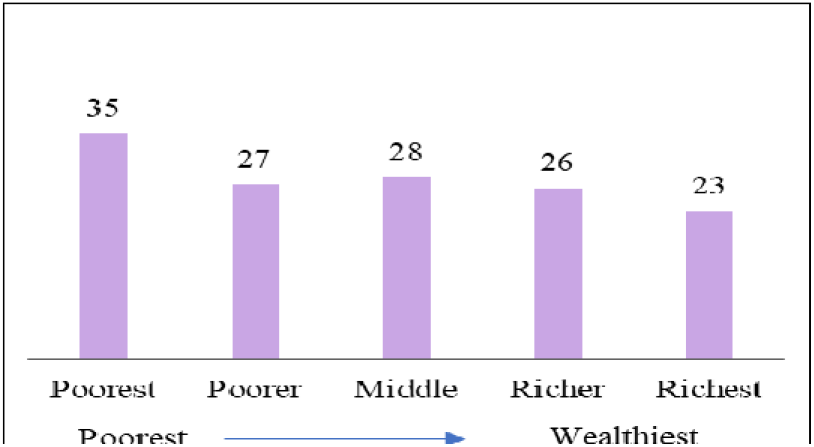
Unintended pregnancy by household wealth

The second strongest predictor was the odds ratio (aOR = 9.56, 95% CI = 6.14–14.89) for currently using any contraception. This implies that respondents who were currently using any contraception were 9.56 times more likely to report unintended pregnancy before their last pregnancy than those who were not currently using any contraception, even when other factors are accounted for in the model. Women between the ages of 15 and 19 had a higher odds ratio (aOR = 1.62, 95% CI = 1.01–2.59) than women in the 35-49 years age group. Unintended pregnancy odds were higher among husbands with primary education (aOR = 1.62, 95% CI = 1.09–2.42) compared to those with no education. It was found that women not desiring any more children (aOR = 2.28, 95% CI = 1.83–2.83) actually had higher odds of unintended pregnancy compared with those who wanted more children. Women who had never terminated a pregnancy were less likely to experience unintended pregnancy (aOR = 0.23, 95% CI = 0.19–0.28) compared to those with a history of termination. The poorest had higher odds (aOR = 1.62, 95% CI = 1.14– 2.29) than the richest (**Table 4**).

**Table 4.** Factor associated with unintended pregnancy among women, NDHS-2022.

| Characteristics | COR* (95% CI) | AOR* (95% CI) | p-value<sup>1</sup> |
| --- | --- | --- | --- |
| **Age groups** |  |  |  |
| 15-19 | 0.51 (0.35-0.73) | 1.62 (1.01- 2.59) | 0.042 |
| 20-24 | 0.38 (0.29-0.50) | 0.93 (0.65-1.32) | 0.699 |
| 25-29 | 0.35 (0.26-0.46) | 0.58 (0.42-0.81) | 0.002 |
| 30-34 | 0.38 (0.28-0.53) | 0.50 (0.35-0.71) | 0.000 |
| 35-49 | Ref. | Ref. |  |
| **Educational attainment** |  |  |  |
| No education | Ref. | Ref. |  |
| Primary | 1.04 (0.80-1.37) | 1.01 (0.72-1.40) | 0.943 |
| Secondary | 0.81 (0.65-1.02) | 0.87 (0.64-1.19) | 0.410 |
| Higher | 0.70 (0.54-0.91) | 0.96 (0.65-1.42) | 0.864 |
| **Husband's education level** |  |  |  |
| No education | Ref. | Ref. |  |
| Primary | 1.58 (1.13-2.21) | 1.62 (1.09-2.42) | 0.017 |
| Secondary | 1.07 (0.79-1.44) | 1.37 (0.93-2.01) | 0.105 |
| Higher | 0.89 (0.64-1.23) | 1.32 (0.84-2.07) | 0.847 |
| **Religion** |  |  |  |
| Hindu | 0.81 (0.58-1.14) | 0.97 (0.66-1.40) | 0.879 |
| Buddhist | 0.54 (0.32-0.91) | 0.61 (0.34-1.07) | 0.087 |
| Muslim | 0.44 (0.26-0.75) | 0.65 (0.35-1.20) | 0.176 |
| Other (Kirat/Christian) | Ref. | Ref. |  |
| **Wealth index** |  |  |  |
| Poorest | 1.79 (1.37-2.33) | 1.62 (1.14-2.29) | 0.006 |
| Poorer | 1.23 (0.93-1.63) | 1.28 (0.90-1.83) | 0.159 |
| Middle | 1.31 (0.99-1.73) | 1.38 (0.98-1.94) | 0.063 |
| Richer | 1.20 (0.91-1.59) | 1.17 (0.84-1.63) | 0.348 |
| Richest | Ref. | Ref. |  |
| **Currently working status** |  |  |  |
| Yes | 1.33 (1.12-1.57) | 0.92 (0.76-1.11) | 0.409 |
| No | Ref. | Ref. |  |
| **Want more children** |  |  |  |
| Yes | Ref. | Ref. |  |
| No | 2.46 (2.06-2.94) | 2.28 (1.83-2.83) | 0.000 |
| **Ever had a terminated pregnancy** |  |  |  |
| Yes | Ref. | Ref. |  |
| No | 0.30 (0.25-0.36) | 0.23 (0.19-0.28) | 0.000 |
| **Currently using any contraception** |  |  |  |
| Yes | 1.38 (1.17-1.62) | 9.56 (6.14-14.89) | 0.000 |
| No | Ref. | Ref. |  |
| **Unmet need** |  |  |  |
| Yes | 1.99 (1.67-2.37) | 14.05 (8.95- 22.07) | 0.000 |
| No | Ref. | Ref. |  |
| * Weighted sample; CI: confidence interval; AOR: adjusted odds ratio; COR: Crude odds ratio; <sup>1</sup>p < 0.05 |  |  |  |

## DISCUSSION

The present study examines the relationship between the unmet need for contraception and unintended pregnancy in Nepal. Despite a mounting trend in contraceptive use among young Nepalese women over the past decade (12), this study found a high occurrence of unintended pregnancy (27.9%) and an unmet need for contraception (29.4%) in Nepal. Results suggest a significant association between unintended pregnancy and an unmet need for contraception. Women with unmet contraceptive needs were significantly associated with unintended pregnancy. This relationship became stronger in the multivariate analysis, indicating that women with unmet contraceptive needs were over 14 times more likely to report unintended pregnancy compared to those without such unmet needs. This finding underscores the critical role of accessible and effective family planning services in preventing unintended pregnancies. Despite national efforts to expand contraceptive coverage, the persistence of unmet need suggests systemic barriers— ranging from supply chain gaps and provider bias to sociocultural constraints—that hinder women’s reproductive autonomy. The magnitude of the association also highlights the urgency of targeted interventions, particularly for populations with limited access to reproductive health services. Addressing unmet need not only reduces unintended pregnancies but also contributes to broader public health goals, including maternal health improvement and gender equity. These results align with global evidence and reinforce the importance of integrating family planning into primary healthcare systems with a rights-based approach, this result also is in line with NDHS and Similar studies conducted in other LMICs like Bangladesh, Ethiopia, and Angola (13–17). Furthermore, links have been established between unintended pregnancies and an elevated risk of abortion and maternal health complications (18). Women burdened with unintended pregnancies are more likely to resort to unsafe abortion practices, which can lead to severe health consequences and potential life-threatening complications (19). So unintended pregnancy and the unmet need for modern contraceptive methods are of considerable concern for women’s reproductive health in Nepal.

The second strongest predictor of reporting unintended pregnancy in Nepal was identified as the current use of any contraception. The study results imply that women who were currently using any form of contraception were almost ten times more likely to report an unintended pregnancy compared to those who were not currently using any contraceptive method. The data accounts for the women’s history before their last pregnancy, indicating a lack of desire for a child at that time, which suggests potential inconsistencies or gaps in contraceptive use leading to unintended pregnancy. Furthermore, the choice of contraceptive method (20), husband’s disapproval or fear of decreased sexual pleasure,(21) the effectiveness of the contraceptive used might have influenced the decision to contraceptive use before their last unintended pregnancy (22). Other significant predictors of reporting unintended pregnancy included age, the husband’s educational status, the desire not to have any more children, and having no history of abortion or terminated pregnancy. This suggests that younger women, particularly those aged 15-19 years, possess a greater likelihood of experiencing unintended pregnancy compared to older women, which is similar to the findings of (15) (23), but does not match with several studies conducted in African settings (24,25). That could be due to a lack of comprehensive knowledge regarding contraceptive methods, combined with restricted access to sexual education stemming from socio-cultural norms, as well as inadequate availability of contraceptive services. Studies in Nepal also showed that religion acts as another contributing factor; Hindu women exhibit a lower likelihood of experiencing unintended pregnancy in this age group-particularly Kirat/Christian women more experiences among non-Hindu (35). Additionally, early marriage or premature initiation of sexual activity can result in a prolonged period of risk for pregnancy, thereby elevating the chances of getting pregnant (23). Identifying preferred contraceptive methods, coupled with the incapacity to negotiate safer sex practices within sexual relationships, might be an additional challenge for this age group. For women whose husbands have a low level of education, the inability to independently make reproductive health decisions(26), coupled with the husband’s disapproval or fear of decreased sexual pleasure (21), can also lead to unintended pregnancies. Women who do not desire to bear any more children demonstrate a greater likelihood of experiencing unintended pregnancy than ones who wish for more offspring. This finding is consistent with studies in Ethiopia, which demonstrated that the lower the desired number of children, the higher the risk of unintended pregnancy (24,25). The reason for this could be that every pregnancy is unintended for a woman who does not want additional children (24). Discordance in fertility intentions—where husbands desire more children than their wives—often results in pressure on women to continue childbearing. When such discordance coincides with inconsistent or non-use of contraceptives, whether due to limited autonomy, inadequate spousal communication, or structural barriers, the likelihood of unintended pregnancy among women who have already achieved their desired family size becomes substantially higher (34). While the past research has explored the negative health effects and contributing factors of unmet contraceptive needs and unintended pregnancies among Nepalese women of reproductive age (27, 28), this study sheds light on the strong relationship between unintended pregnancy and unmet contraceptive needs. The elevated unintended pregnancy rate in Karnali (38%) aligns with prior findings that remote regions face systemic barriers, including limited healthcare infrastructure and lower contraceptive uptake (10). Targeted interventions, such as mobile clinics and culturally tailored counseling, may help reduce this disparity (30). These regional variations underscore the need for geographically sensitive reproductive health strategies. Provinces with higher rates of unintended pregnancy and completed fertility preferences may benefit from expanded access to long-term contraceptive methods and counseling services. Meanwhile, regions with greater demand for birth spacing require robust support for short-term methods and community-based education. Addressing these disparities is essential for improving reproductive autonomy and aligning Nepal’s family planning efforts with its FP-2030 commitments and Sustainable Development Goals (17, 31, 32). This research adds to the expanding knowledge base by confirming that various factors such as age, husband’s education level, women’s autonomy, desire to have more children and spousal dynamics significantly influence contraceptive practices and unintended pregnancy rates. The significant association between unmet contraceptive needs and unintended pregnancy underscores the importance of addressing gaps in access to and consistent use of contraceptive methods.

## CONCLUSION

This study underscores the strong link between unmet contraceptive needs and unintended pregnancies among married women in Nepal. Women with unmet need were significantly more likely to report unintended pregnancies, even after adjusting for socio-demographic and reproductive factors. The analysis also revealed that younger age, lower household wealth, limited education, and completed fertility preferences were associated with higher odds of unintended pregnancy. These findings highlight the importance of improving access to contraceptive services and tailoring interventions to the needs of vulnerable subgroups. Addressing gaps in contraceptive coverage and promoting informed reproductive choices are essential for reducing unintended pregnancies and their associated health risks. A comprehensive approach that integrates individual, relational, and systemic factors is vital to advancing reproductive health outcomes in Nepal.

### Limitations

Several limitations should be acknowledged when interpreting the findings of this study. First, the cross-sectional design restricts the ability to draw causal inferences between unmet contraceptive needs and unintended pregnancy. Second, key variables such as pregnancy intention and contraceptive needs were self-reported by participants, which may be subject to recall bias or influenced by social desirability, potentially affecting the accuracy of the data. Finally, the study did not incorporate qualitative approaches to explore deeper cultural, behavioral, or interpersonal factors that might shape contraceptive use and pregnancy planning, limiting understanding of the complex social dynamics underlying these outcomes.

### Recommendation

To reduce unintended pregnancies and address unmet contraceptive needs, several strategies should be prioritized. First, culturally responsive counseling programs that align with local beliefs, norms, and misconceptions are essential; community health workers can play a vital role by engaging families to address concerns about side effects and religious taboos related to contraceptive use. Second, reproductive health education should be integrated into school curricula and youth platforms, with a special focus on adolescents aged 15–19, who face higher risks of unintended pregnancy. Third, promoting male involvement through couple-based counseling can encourage shared decision-making and help dispel myths about contraception’s impact on sexual satisfaction. Fourth, ensuring consistent availability of modern contraceptive methods in both urban and rural health facilities is critical, alongside outreach services that reach remote and underserved communities. Finally, empowering women to make independent reproductive choices through education, economic opportunities, and health literacy initiatives will strengthen their autonomy and improve reproductive health outcomes.

### Ethics approval

The analysis of this study is based on this publicly available dataset. We obtained permission from the DHS Program to use the NDHS 2022 dataset, which can be downloaded from https://www.dhsprogram.com. The NDHS 2022 received ethical approval from both the Institutional Review Board of ICF International, USA (Reference number: 180657.0.001.NP.DHS.01, Date: April 28, 2022), and the Ethical Review Board of the Nepal Health Research Council (Reference number: 678, Date: September 30, 2021). The NDHS 2022 maintained written informed consent from all adult participants, and consent or assent was obtained from parents or guardians for participants under the age of 18.

## Acknowledgments

The authors are thankful to the USAID Demographic and Health Surveys (DHS) Program for granting access NDHS 2022 dataset. icddr,b is thankful to the Governments of Bangladesh and Canada for providing unrestricted/institutional support.

## Authors’ contributions

AAS, TW, KIT, MM, OA, and MGR conceptualized the research idea, analyzed the data, and wrote the manuscript under SSM’s supervision. All authors read and approved the final manuscript.

## Data availability

Permission to use the NDHS 2022 dataset was granted by the DHS Program, and the dataset is available for download at https://www.dhsprogram.com.

